# Community vaccination can shorten the COVID-19 isolation period: an individual-based modeling approach

**DOI:** 10.1101/2022.02.08.22270668

**Authors:** Chayanin Sararat, Jidchanok Wangkanai, Chaiwat Wilasang, Tanakorn Chantanasaro, Charin Modchang

## Abstract

**Background:** Isolation of infected individuals and quarantine of their contacts are usually employed to mitigate the transmission of SARS-CoV-2. While 14-day isolation of infected individuals could effectively reduce the risk of subsequence transmission, it also significantly impacts the patient’s financial, psychological, and emotional well-being. It is, therefore, vital to investigate how the isolation duration could be shortened when effective vaccines are available and in what circumstances we can live with COVID-19 without isolation and quarantine.

**Methods:** An individual-based modeling approach was employed to estimate the likelihood of secondary infections and the likelihood of an outbreak following the isolation of an index case for a range of isolation periods. Our individual-based model integrates the viral loads and infectiousness profiles of vaccinated and unvaccinated infected individuals. The effects of waning vaccine-induced immunity against Delta and Omicron variant transmission were also investigated.

**Results:** In the baseline scenario in which all individuals are unvaccinated, and no nonpharmaceutical interventions are employed, there is a chance of about 3% that an unvaccinated index case will make at least one secondary infection after being isolated for 14 days, and a sustained chain of transmission can occur with a chance of less than 1%. We found that at the outbreak risk equivalent to that of 14-day isolation in the baseline scenario, the isolation duration can be shortened to 7.33 days (95% CI 6.68-7.98) if 75% of people in the community are fully vaccinated during the last three months. In the best-case scenario in which all individuals in the community are fully vaccinated, isolation of infected individuals may no longer be necessary. However, to keep the outbreak risk low, a booster vaccination may be necessary three months after full vaccination. Finally, our simulations showed that the reduced vaccine effectiveness against transmission of the Omicron variant does not much affect the risk of an outbreak if the vaccine effectiveness against infection is maintained at a high level via booster vaccination.

**Conclusions:** The isolation duration of a vaccine breakthrough infector could be safely shortened if a majority of people in the community are immune to SARS-CoV-2 infection. A booster vaccination may be necessary three months after full vaccination to keep the outbreak risk low.

## Background

SARS-CoV-2 spreads rapidly throughout the world, causing over 288.23 million infections and 5.48 million deaths by the end of 2021 [1]. During the early phase of transmission, when vaccines were unavailable, nonpharmaceutical interventions have been frontline measures to mitigate the transmission [2, 3]. Isolation of infected individuals is a critical strategy widely employed to break the transmission chain. Institution-based isolation of confirmed cases has been shown in a modeling study to delay the epidemic’s peak and reduce the epidemic’s size by approximately 57% [4]. Isolation, however, will be effective only if it can be promptly employed to prevent pre-symptomatic and asymptomatic transmission [5]. In addition, the isolation period should also be long enough to ensure that the infected individuals do not spread the disease after the isolation. However, while prolonged isolation may reduce the risk of transmission more effectively, it also significantly impacts the patient’s financial, psychological, and emotional well-being [6-8].

COVID-19 vaccines were first made available in the last of 2020 [9], and they have been shown to be effective at preventing infection and transmission [10-12]. Despite the fact that infections can occur even after being fully vaccinated, a faster viral clearance was observed in the breakthrough infections, indicating that a breakthrough infected individual may have a shorter duration of infectiousness [13, 14]. As a result, it suggests that those who have been vaccinated may require a shorter period of isolation. It is vital to comprehend how the isolation period could be reduced based on vaccine effectiveness, particularly when we desire to return to normalcy and live with COVID-19 without quarantine and isolation measures.

In this study, we used an individual-based modeling approach to assess the likelihood of secondary infections and the likelihood of an outbreak following isolation of a vaccinated index case for a range of isolation periods. Our individual-based model accounts for transmission heterogeneity, variation in the course of infection, and the disease’s infectivity profiles of both vaccinated and unvaccinated infected individuals. The effects of waning vaccine-induced immunity and the delay in isolating infected individuals in the community were also examined.

## Methods

### Estimation of infectiousness profiles and vaccine efficiency against transmission

Individuals infected with SARS-CoV-2 can become infectious prior to the onset of symptoms. In the case of unvaccinated individuals, the infectiousness peaks 2.1 days before the onset of symptoms and then decreases gradually during the course of the illness [15]. Although the viral trajectories during the proliferation stage are similar in both unvaccinated and vaccinated individuals, the viral loads are cleared faster in vaccine breakthrough infections than in the unvaccinated individuals [13, 14]. The disease infectiousness (*F*) prior to the peak of both vaccinated and unvaccinated infectors was therefore assumed to follow a gamma distribution, as described in [15]. After the peak period, data on *C*_*t*_ values collected in Singapore [13] were used to determine the infectiousness profile. The infectiousness was considered to be directly proportional to the viral load (*V*) that exceeds a threshold of 10^6^ copies, i.e., *F∝Vx10*^*-6*^ [16]. *C*_*t*_ values were converted to viral load (*V*) using the procedures outlined in [14]. Because of the faster viral clearance time, the disease transmissibility of vaccinated infectors could be averted compared to unvaccinated ones. In this work, we estimated the vaccine efficacy against transmission from a percentage of reduction in the area under the curves of the disease infectiousness profiles of vaccinated and unvaccinated infectors.

### Model structure

In order to examine the probability of post-isolation infections, the transmissions of the COVID-19 were simulated with individuals categorized as susceptible (*S*), latent (*L*), infectious (*I*), recovered (*R*), isolated (*Q*), and fully vaccinated (*V*) according to their infection and vaccination status. Infectious individuals are further divided into symptomatic (*I*_*S*_) and asymptomatic (*I*_*A*_) infectious individuals, with the assumption that asymptomatic infectious individuals are less infectious than symptomatic ones. Although COVID-19 vaccines cannot entirely protect people against infection, they are still beneficial in decreasing the chance of infection. In addition, even when vaccinated individuals get infected, they will be less likely to transmit the disease to other individuals. In our model, vaccine breakthrough infections are distinguished from infections in susceptible individuals by subscripts *V* and *S*, as shown in **Figure 1(A)**. After being infected, individuals enter a latent state before becoming infectious. Finally, infectious individuals move either to recovered or isolated compartments.

**Figure 1:**
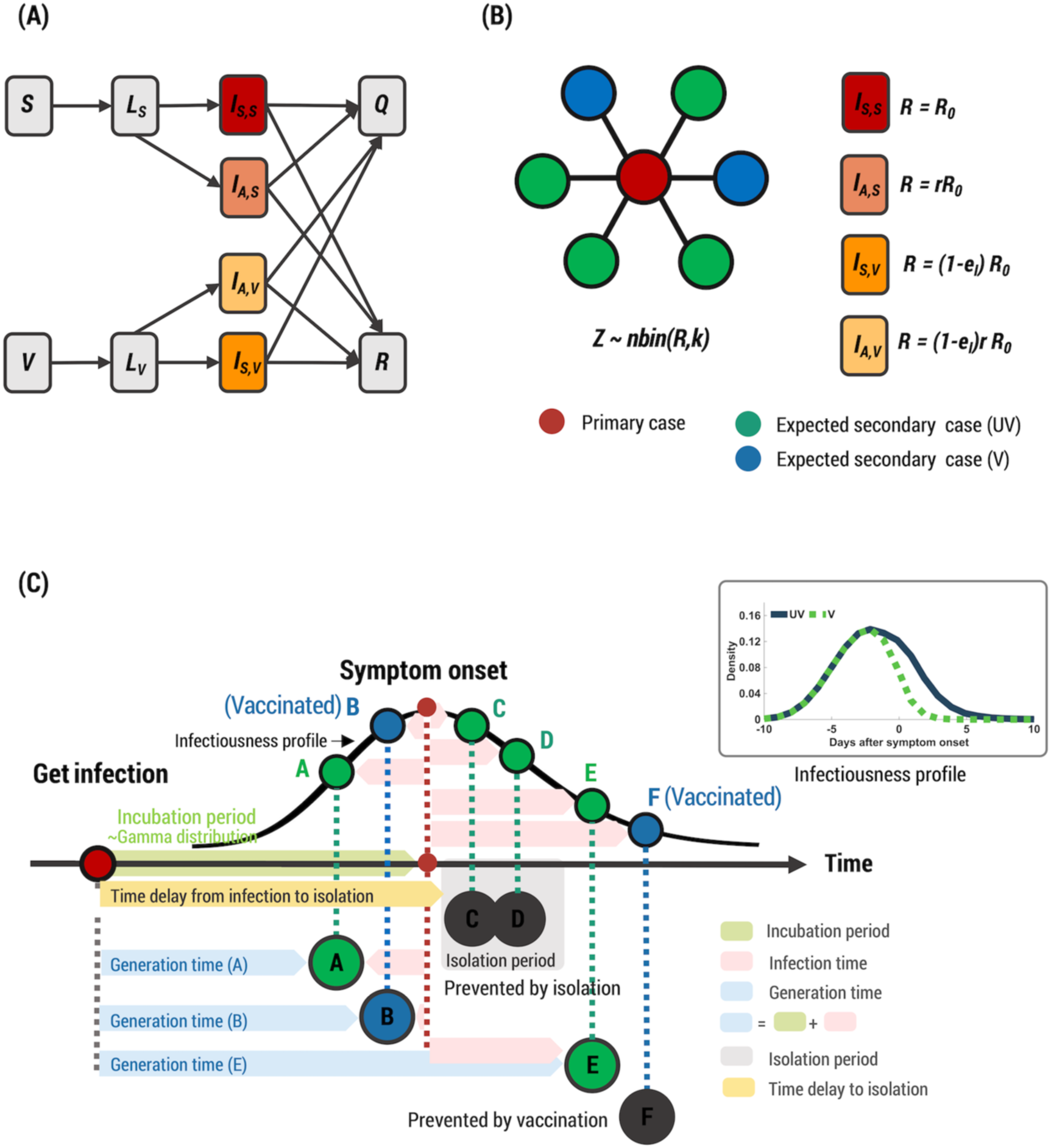
Model structure of the COVID-19 transmission. **(A)** Schematic of the compartmental model showing progressive of the disease and transition of individuals across different compartments. **(B)** Example of the expected number of secondary cases made by a single primary case drawn from a negative binomial distribution. **(C)** An illustration of transmission events due to the primary case (red circle). The generation time is a time duration between a primary case’s infection and one of its subsequent secondary cases. The incubation period is a time duration from exposure to symptom onset. The inset shows the infectiousness profiles of unvaccinated (UV) and vaccinated (V) infected individuals.

The number of secondary infections caused by a single primary case, *Z*, for each infected individual is estimated from a negative binomial distribution with a mean equal to the reproduction number (*R*_*0*_) and dispersion parameter (*k*) (**Figure 1(B))**. Because of the lower infectivity of asymptomatic infectious individuals, they contribute fewer infections; the mean number of secondary cases made by an asymptomatic infectious individual was reduced by a factor *r*. For the vaccine breakthrough infectors, the mean of the distribution is also reduced due to the efficiency against transmission of vaccines (*e*_*I*_).

An example of the transmission events is illustrated in **Figure 1(C)**. The incubation period, time from exposure to symptom onset, is assumed to follow the Gamma distribution with a mean of 5.8 days [15]. The time of each new infection is drawn from a random number distribution that is distributed according to the infectiousness profile of the infectors. As a result of the vaccine’s effectiveness against infection, vaccinated individuals are less likely to become infected. The effective infectious period is determined by whether or not infected individuals are isolated. If infected individuals are isolated, they will be contagious until they are isolated. Although transmission can be prevented during isolation, post-isolation infections are still possible. The generation time between infection of a primary case and one of its subsequent secondary cases is dependent on both the incubation period and the infection time. In our study, the primary index case is assumed to be isolated immediately after becoming infected, whereas other subsequent infected individuals in the community are isolated with a default time delay of 6.8 days.

### Estimating the probability of secondary transmission and the probability of a successful outbreak after isolation

The probability of secondary transmission was defined as a chance that a primary case makes at least one subsequence infection after the isolation. The probability of a successful outbreak was estimated from the likelihood that the chain of transmission initiated from the index case after isolation continues for more than 90 days. We have checked that the threshold value of 90 days can distinguish between simulations in which the disease goes extinct and simulations in which the disease spreads substantially until reaching the equilibrium state. The probabilities were estimated using three batches of simulations, each containing 1,000 realizations.

**Table 1:**
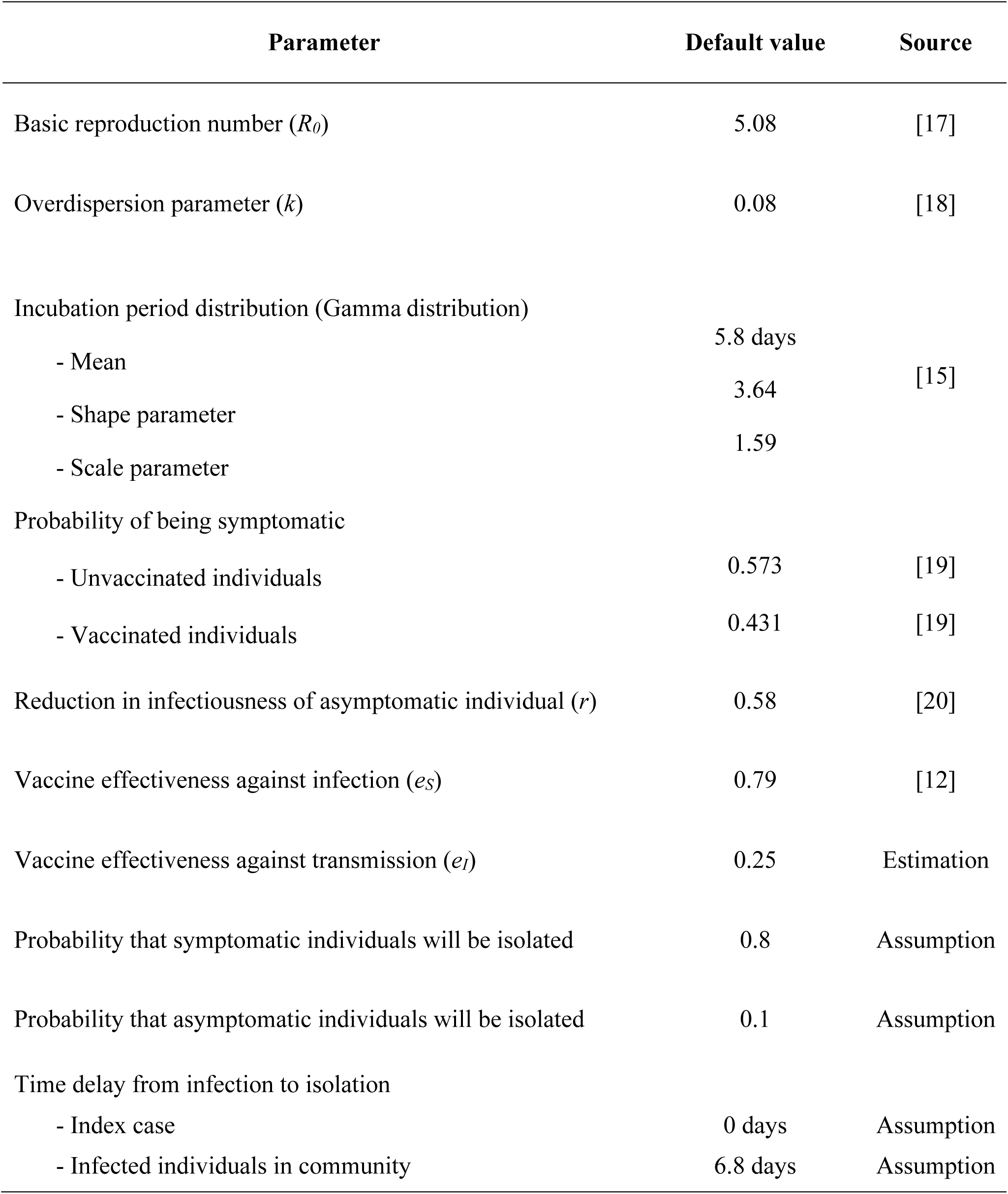
Model parameters and their default values.

## Results

### Impact of vaccination on post-isolation transmission

We explored the probability of a primary infected individual making at least one secondary infection and the probability of a successful outbreak, i.e., having a sustained chain of transmission, after being released from isolation. In the baseline scenario in which the primary case and all other individuals in the community are unvaccinated, we found that there is a chance of about 3% that the unvaccinated index case will make at least one secondary infection after being isolated for 14 days, and a sustained chain of transmission can occur with a chance of less than 1% (left bars in **Figure 2(A)-(B)**). However, if the index case has already been vaccinated, we found that although all other individuals in the community are unvaccinated, only about 10 days of isolation is equivalent to 14-day isolation of unvaccinated index case (red lines and red symbols in **Figure 2**).

**Figure 2:**
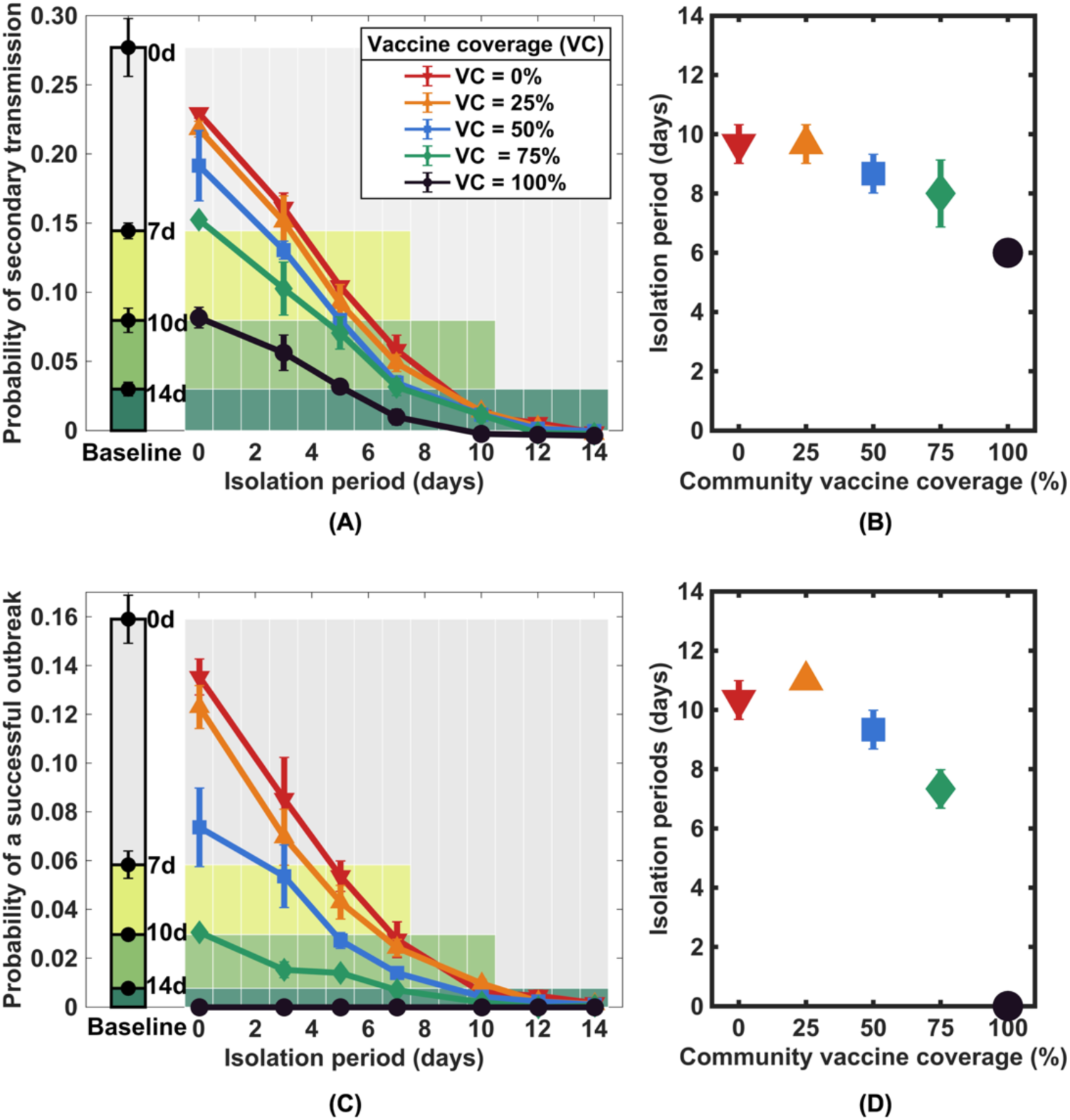
Impacts of isolating a primary vaccinated infector on post-isolation transmission. Probability of secondary transmission **(A)** and probability of a successful outbreak in which a chain of transmission can be sustained **(C)** after a range of isolation periods and vaccination levels in the community. The corresponding probabilities in the baseline scenario where the index case and all other individuals in the community are unvaccinated are shown as bar graphs on the left side of both subfigures. **(B)** and **(D)** show the isolation period equivalent to the 14-day isolation period in the baseline scenarios, regarding the probability of secondary transmission and the probability of a successful outbreak, respectively. Error bars indicate 95% CI.

Vaccinating people in the community can further reduce the likelihood of secondary infections and the probability of a successful outbreak. It was found that higher community vaccine coverages decrease the chance of secondary transmission following the isolation of the vaccinated index case more, especially when the isolation periods are short. In addition, when the isolation period is longer than 12 days, there is no apparent difference between different vaccination coverages. At the outbreak risk equivalent to that of 14-day isolation in the baseline scenario, the isolation duration of the primary vaccinated infector can be shortened to 9.33 days (95% CI 8.68-9.98) if 50% of people in the community are vaccinated (**Figure 2 (D)**). When 75% of people in the community are vaccinated, the isolation period can be further shortened to 7.33 days (95% CI 6.68-7.98). Finally, we found that in the best-case scenario in which all individuals are vaccinated, although post-isolation infections are still possible for the isolation period of shorter than 6 days, the chance of sustained chain of transmission to occur is extremely rare. In this case, isolation of infected individuals may no longer be necessary.

### Effect of waning vaccine-induced immunity

As the vaccine effectiveness against Delta variant infection decreases over time [21], we evaluated its effect on the probability of secondary infections and the probability of a successful outbreak following isolation. We found that for a low level of immunization (< 25% coverage), both the post-isolation transmission probability and the successful outbreak probability are not significantly affected by the waning of vaccine effectiveness (**Figure 3 (A)** and **(D)**). However, for higher vaccine coverage, the effect of the decline in the vaccine effectiveness is more pronounced, especially when the isolation durations are short. Note, however, that although at high vaccination coverage (> 75% coverage), there is a more significant effect of immunity waning across a range of isolation periods, the probability of an outbreak is still lower than that in the case when 25% of the population are vaccinated. With the vaccine coverage of 75%, for example, after four months of vaccination, the outbreak risk climbs from 0.9% to 4.2% for 3-day isolation and increases from 1.3% to 7.7% for no isolation (**Figure 3 (E)**). When all individuals in the community are vaccinated, despite a substantial decrease in vaccine effectiveness after four months, the chance of a successful outbreak is still lower than 4% even there is no isolation (**Figure 3 (F)**).

**Figure 3:**
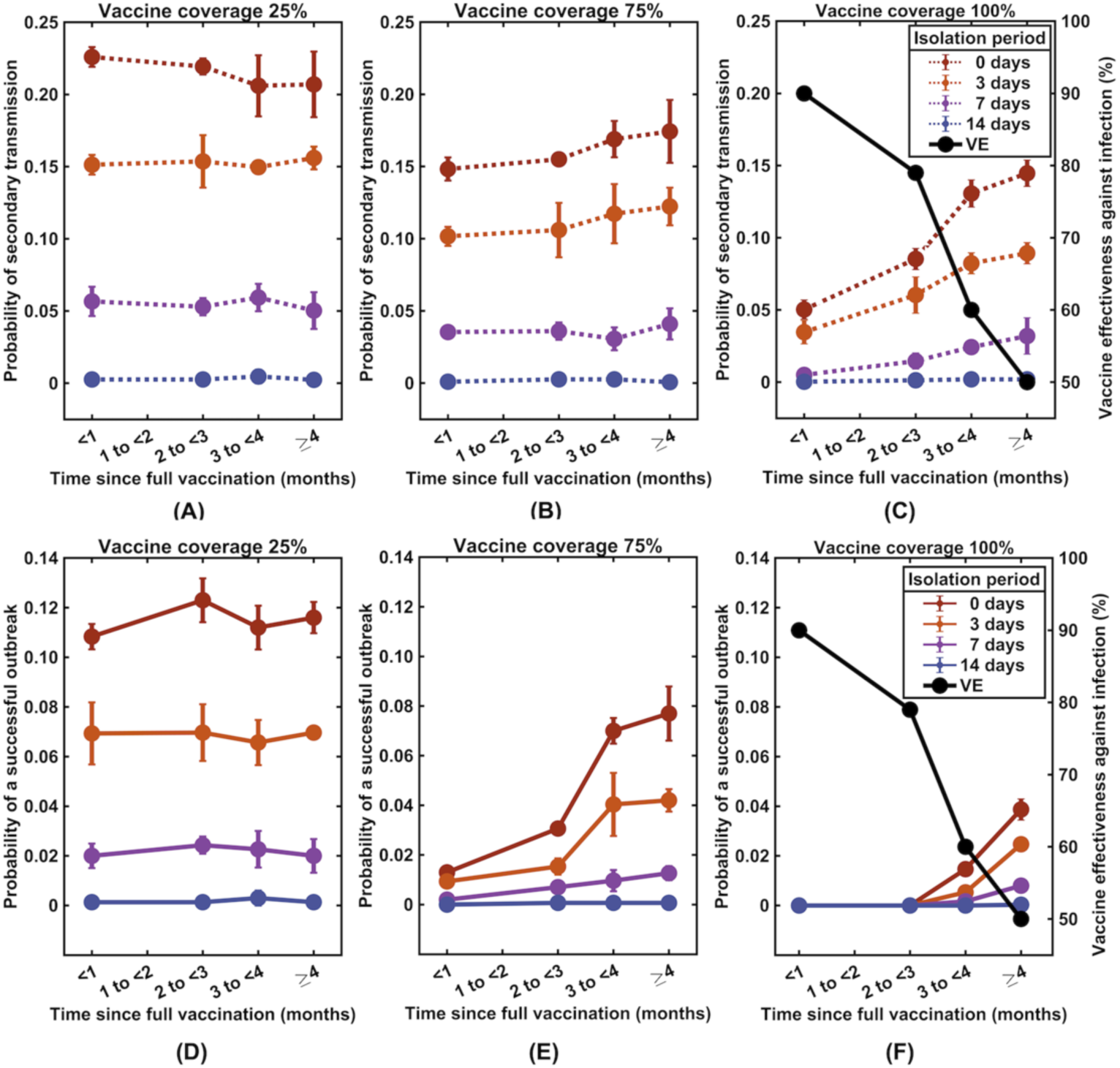
The effect of reduction in vaccine effectiveness against SARS-CoV-2 infection. The time evolution of the probability of at least one secondary infection **(A-C)** and probability of a successful outbreak **(D-F)** following the release of a breakthrough infector from isolation as the vaccine effectiveness against infection wanes (black lines, right y-axis). Data of vaccine effectiveness against infection were obtained from reference [21].

We also investigated how the change in vaccine effectiveness against transmission would influence the likelihood of secondary infections and the probability of a successful outbreak. In this part, we considered the vaccine effectiveness against transmission (*e*_*I*_) ranges from 0% to 40%, and the vaccine effectiveness against infection (*e*_*S*_) of 90% and 50%, corresponding to the effectiveness against infection of Delta variant after being fully vaccinated with two doses of mRNA BNT162b2 vaccine for one month and four months, respectively. We found that during the first four months after complete vaccination, when the vaccine effectiveness against infection is high, the vaccine effectiveness against transmission had only a minor effect on the transmission, especially when the isolation period is long (**Figure 4**).

**Figure 4:**
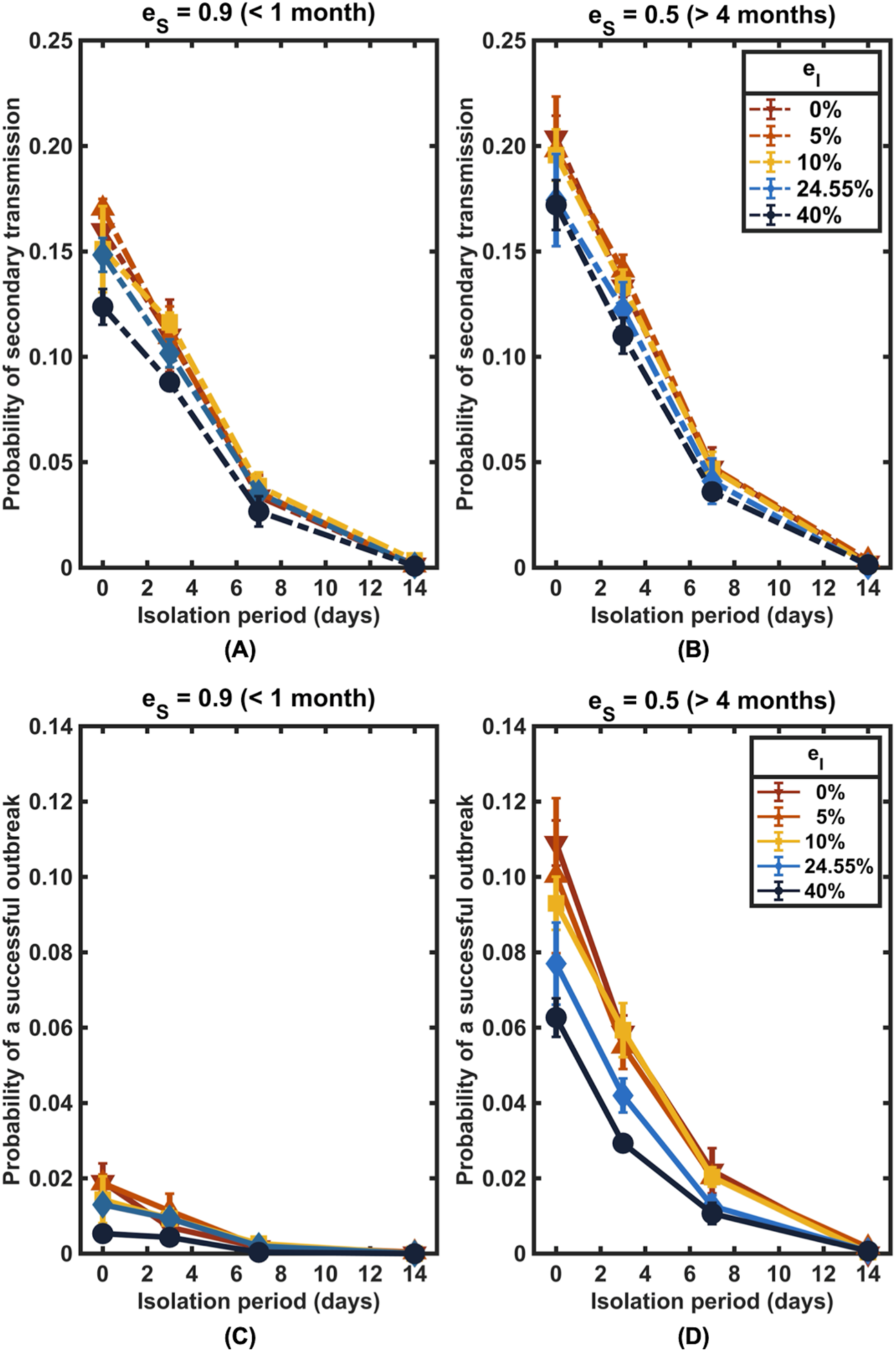
The influence of vaccine effectiveness against SARS-CoV-2 transmission. The probability of at least one secondary infection (**(A)** and **(B))** and a successful outbreak (**(C)** and **(D))** after being released from isolation into a community with a vaccination level of 75%. The vaccine effectiveness against transmission (*e*_*I*_) was varied from 0% to 40%, and the vaccine effectiveness against infection (*e*_*S*_) was fixed at 90% (left column) and 50% (right column).

### Impact of community case-isolation and other control measures

We next evaluated the impact of time delay from infection to the isolation of infected individuals in the community on the spread of SARS*-*CoV*-*2. Our results indicated that the outbreak would be less likely to occur if case isolation is performed with a shorter delay (**Figure 5**). For example, under the vaccine coverage of 75%, the outbreak risk could be suppressed to lower than 1% if the isolation can be performed within 3 days after infections. To maintain the same level of an outbreak risk, a longer duration of the isolation is needed for the isolation with longer delays. For instance, for a 5-day delay, at least 5 days of isolation may be required, and for a 7-day delay, at least 7 days of isolation may be needed. When only 25% of individuals are vaccinated, isolation may be required for at least 10 days, regardless of how quickly infected individuals are isolated.

**Figure 5:**
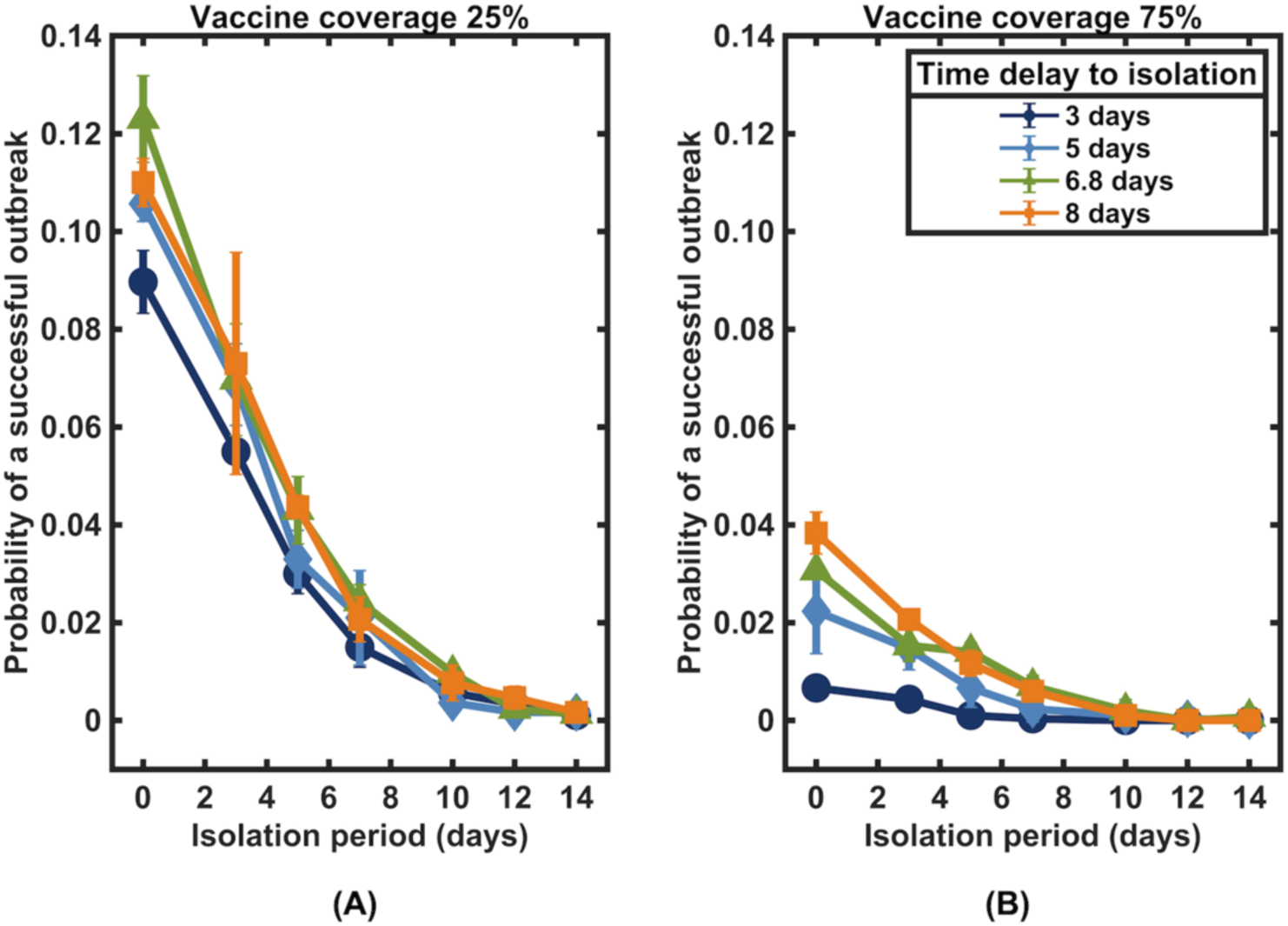
Impact of time delay from infection to isolation under vaccination coverages of (A) 25% and (B) 75%.

The effective reproduction number (*R*) is commonly used to measure the disease transmissivity under different control measures. To consider the effects of other control measures, a sensitivity analysis on the effective reproduction number has been performed. In combination with other non-pharmaceutical interventions, we found that community vaccination could further shorten the isolation period (**Figure 6**). For instance, in the absence of any non-pharmaceutical interventions and the vaccine coverage is only 25%, case isolation may be required for at least 12 days to reduce the outbreak risk to below 1%. However, if other control measures are concurrently implemented at a level that could reduce the effective reproduction number to 3.2, only one week of isolation is sufficient. Importantly, in this case, isolation will be no longer necessary if the community vaccination level reaches 75%.

**Figure 6:**
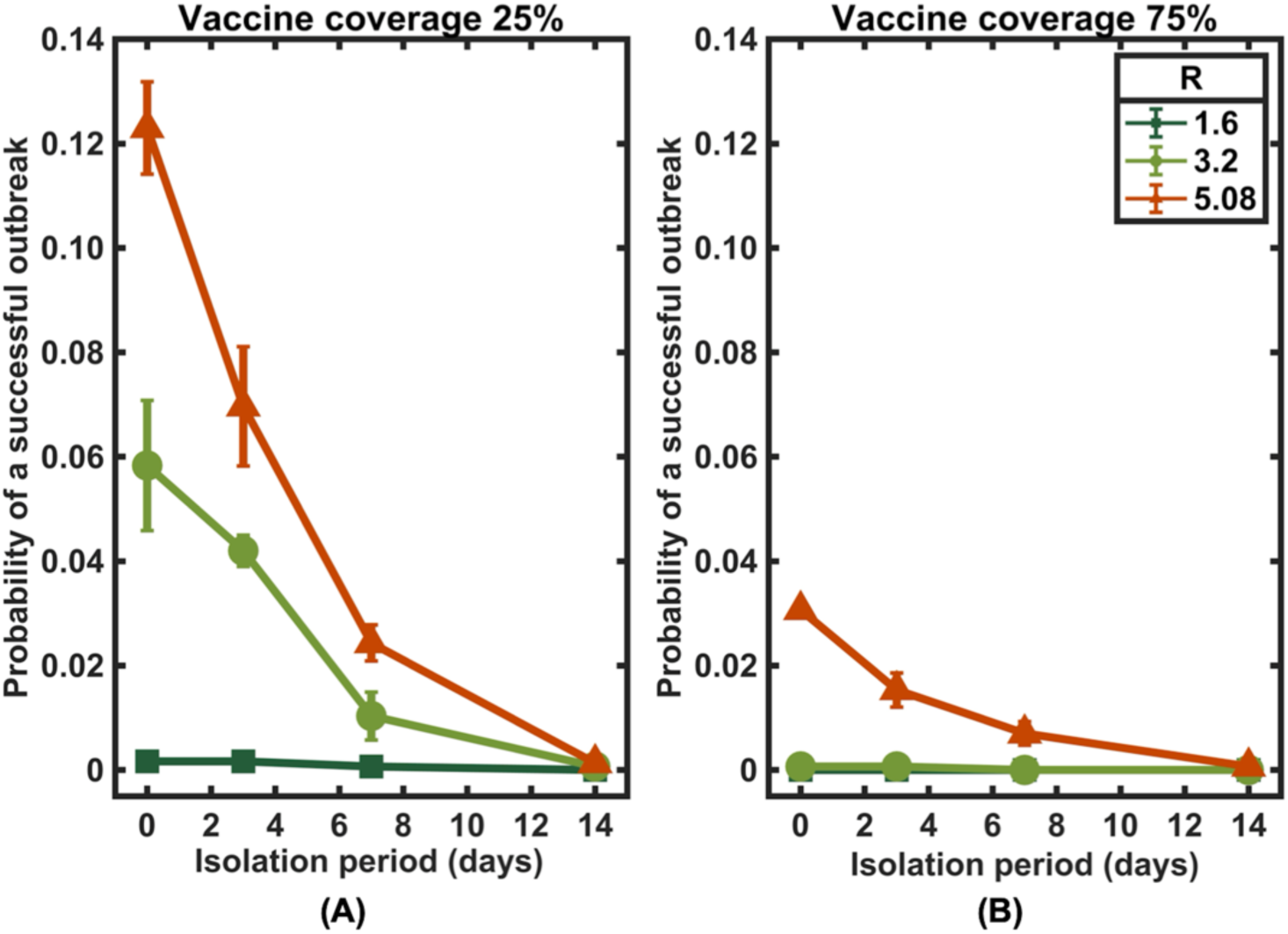
A sensitivity analysis on the effective reproduction number. The probability of a successful outbreak under the community vaccination coverages of **(A)** 25% and **(B)** 75%.

## Discussion

In this work, we evaluated the likelihood of at least one secondary infection and the likelihood of an outbreak following the isolation of a vaccine breakthrough infector for a specified period of time. Our modeling results indicated that vaccines play a critical role in reducing the likelihood of post-isolation transmission. We discovered that the duration of isolation for an infected individual who has already been vaccinated could be reduced as opposed to the 14-day duration of isolation for unvaccinated individuals. Additionally, the duration of isolation can be reduced further if the majority of the community members are immune to the disease (**Figure 2**).

In the best-case scenario in which all individuals in the community are fully vaccinated with two doses of mRNA BNT162b2 vaccine, isolation of Delta-variant breakthrough infected individuals may no longer be required, at least during the first three months after being fully vaccinated, if no other non-pharmaceutical interventions are implemented. After three months, however, as the vaccine effectiveness against infection drops to around 60% [21], the probability of post-isolation transmission increases rapidly after this time, especially in the cases of short isolation periods. This result indicates that booster vaccination may be needed after being fully vaccinated for three months; otherwise, more extended isolation periods or other non-pharmaceutical control measures may be necessary to compensate for the increased transmission risk (**Figure 3**).

With a faster viral clearance time in vaccinated individuals, vaccines have been hypothesized to reduce onward transmission from infected vaccinated individuals. According to our estimations, the vaccine effectiveness against transmission of 24.6% is comparable to the effectiveness against transmission with the Delta variant after getting two doses of the Pfizer vaccine [22]. However, the emergence of the Omicron variant has raised serious concerns about its capability to evade vaccine protection. After receiving two doses of mRNA vaccines, the vaccine effectiveness in preventing Omicron-variant transmission drops to less than 5% [22]. Nevertheless, our simulations showed that the reduced vaccine effectiveness against Omicron-variant transmission does not much affect the risk of secondary infection if the vaccine effectiveness against infection is restored to a high level via booster vaccination [23] (**Figure 4**). However, since at the time of writing this manuscript, the data on the waning of vaccine effectiveness against Omicron-variant infections were not available, we assumed that they wane at the same rate as the Delta variant.

When considering the effect of delay in isolation of infected individuals in the community, we found that a shorter delay to isolation can further shorten the isolation period, especially in the high vaccine coverage settings. In addition, we found that while an outbreak may still occur in the absence of isolation in the community with low vaccination coverage, the risk could be minimized when additional control measures such as contact tracing and quarantine of their contacts, as well as testing, are implemented (**Figures 5 and 6**).

## Data Availability

All data produced in the present study are available upon reasonable request to the authors.

## Author contributions

CM conceived and designed the study. CS, JW, CW, and TC conducted the study. CS and CM analysed the results. CS and CM wrote the manuscript.

## References

1. Hannah Ritchie, E.M., Lucas Rodés-Guirao, Cameron Appel, Charlie Giattino, Esteban Ortiz-Ospina, Joe Hasell, Bobbie Macdonald, Diana Beltekian and Max Roser, Coronavirus Pandemic (COVID-19). Our World in Data, 2020.

2. Wilasang, C., et al., Reduction in effective reproduction number of COVID-19 is higher in countries employing active case detection with prompt isolation. Journal of travel medicine, 2020. 27(5): p. taaa095.

3. Wilasang, C., et al., Reconstruction of the transmission dynamics of the first COVID-19 epidemic wave in Thailand. Scientific Reports, 2022. 12(1): p. 2002.

4. Dickens, B.L., et al., Institutional, not home-based, isolation could contain the COVID-19 outbreak. The Lancet, 2020. 395(10236): p. 1541–1542.

5. Moghadas, S.M., et al., The implications of silent transmission for the control of COVID-19 outbreaks. Proceedings of the National Academy of Sciences, 2020. 117(30): p. 17513–17515.

6. Pietrabissa, G. and S.G. Simpson, Psychological Consequences of Social Isolation During COVID-19 Outbreak. Frontiers in Psychology, 2020. 11(2201).

7. Jain, A., et al., Impact on mental health by “Living in Isolation and Quarantine” during COVID-19 pandemic. Journal of family medicine and primary care, 2020. 9(10): p. 5415–5418.

8. Hossain, M.M., A. Sultana, and N. Purohit, Mental health outcomes of quarantine and isolation for infection prevention: a systematic umbrella review of the global evidence. Epidemiology and health, 2020. 42: p. e2020038–e2020038.

9. Baraniuk, C., Covid-19: How the UK vaccine rollout delivered success, so far. bmj, 2021. 372.

10. Lopez Bernal, J., et al., Effectiveness of Covid-19 vaccines against the B. 1.617. 2 (Delta) variant. N Engl J Med, 2021: p. 585–594.

11. Harris, R.J., et al., Effect of Vaccination on Household Transmission of SARS-CoV-2 in England. New England Journal of Medicine, 2021. 385(8): p. 759–760.

12. Sheikh, A., et al., SARS-CoV-2 Delta VOC in Scotland: demographics, risk of hospital admission, and vaccine effectiveness. The Lancet, 2021. 397(10293): p. 2461–2462.

13. Chia, P.Y., et al., Virological and serological kinetics of SARS-CoV-2 Delta variant vaccine-breakthrough infections: a multi-center cohort study. medRxiv, 2021: p. 2021.07.28.21261295.

14. Kissler, S.M., et al., Viral dynamics of SARS-CoV-2 variants in vaccinated and unvaccinated individuals. medRxiv, 2021: p. 2021.02.16.21251535.

15. Kang, M., et al., Transmission dynamics and epidemiological characteristics of Delta variant infections in China. medRxiv, 2021: p. 2021.08.12.21261991.

16. Larremore, D.B., et al., Test sensitivity is secondary to frequency and turnaround time for COVID-19 screening. Science advances, 2021. 7(1): p. eabd5393.

17. Liu, Y. and J. Rocklöv, The reproductive number of the Delta variant of SARS-CoV-2 is far higher compared to the ancestral SARS-CoV-2 virus. Journal of Travel Medicine, 2021. 28(7).

18. Endo, A., et al., Estimating the overdispersion in COVID-19 transmission using outbreak sizes outside China [version 3; peer review: 2 approved]. Wellcome Open Research, 2020. 5(67).

19. Tang, L., et al., Asymptomatic and Symptomatic SARS-CoV-2 Infections After BNT162b2 Vaccination in a Routinely Screened Workforce. JAMA, 2021. 325(24): p. 2500–2502.

20. Byambasuren, O., et al., Estimating the extent of asymptomatic COVID-19 and its potential for community transmission: systematic review and meta-analysis. Official Journal of the Association of Medical Microbiology and Infectious Disease Canada, 2020. 5(4): p. 223–234.

21. Tartof, S.Y., et al., Effectiveness of mRNA BNT162b2 COVID-19 vaccine up to 6 months in a large integrated health system in the USA: a retrospective cohort study. The Lancet, 2021. 398(10309): p. 1407–1416.

22. Gardner, B.J. and A.M. Kilpatrick, Estimates of reduced vaccine effectiveness against hospitalization, infection, transmission and symptomatic disease of a new SARS-CoV-2 variant, Omicron (B.1.1.529), using neutralizing antibody titers. medRxiv, 2021: p. 2021.12.10.21267594.

23. Andrews, N., et al., Effectiveness of COVID-19 vaccines against the Omicron (B. 1.1. 529) variant of concern. MedRxiv, 2021.

